# The independent and joint effects of outdoor air pollution exposure and genetic risk on mental health trajectories during adolescence

**DOI:** 10.64898/2026.07.12.26357864

**Authors:** Giulia Cattarinussi, Yingzhe Zhang, Paola Dazzan, Divyangana Rakesh

## Abstract

Air pollution exposure has been associated with increased risk of developing mental health problems. It is possible that individuals at high genetic risk for psychopathology may be more vulnerable to these effects; however, this question remains to be investigated. We leveraged longitudinal data from n=10,620 participants from the Adolescent Brain Cognitive Development Study to first investigate sex-stratified associations of particulate matter (PM2.5) exposure and genetic risk with mental health trajectories across 9-16 years including internalizing symptoms and psychotic like experiences (PLEs). Additionally, we tested whether genetic risk for schizophrenia (PRS-SCZ) and major depressive disorder (PRS-MDD) exacerbate the association with PM2.5 exposure and change in symptoms over time. PM2.5 exposure was associated with lower decreases in PLEs over time in females (p-FDR=0.005), with no effects on internalising symptom trajectories in either sex. Genetic influences were sex-specific, with higher PRS-SCZ and PRS-MDD linked to greater increases in internalising symptoms in females (p-FDR=0.009; p-FDR=0.022) and higher PRS-MDD associated with greater decreases in PLEs in males (p-FDR=0.001). In females we also observed an interaction between PM2.5 and PRS-MDD on PLEs trajectories (p-FDR=0.048) such that those with high genetic risk and high PM2.5 exposure demonstrated increases in PLEs over time. Our results suggest that PM2.5 exposure and polygenic risk for depression jointly shape mental health during adolescence. This underscores the potential of interventions aimed at lowering air pollution during sensitive periods of neurodevelopment in improving adolescent mental health.

## Introduction

Air pollution is a major health issue worldwide, linked to a range of physical health problems, including cardiovascular and respiratory disease (Brunekreef and Holgate, 2002). Emerging evidence also suggests an association between air pollution and higher levels of psychopathology (Bhui et al., 2023). Adolescence represents a particularly sensitive period of development during which many mental health problems, including internalizing symptoms and psychotic like experiences, first emerge (Paus et al., 2008; Solmi et al., 2021). Environmental exposures, including air pollution, may shape the emergence and progression of these symptoms (Hobbs et al., 2025; Newbury et al., 2019). However, the extent to which air pollution contributes to the development and course of psychopathology during this period and whether certain individuals are more susceptible to its effects remains unclear. Given that mental health problems contribute significantly to global burden of disease in youth (Whiteford et al., 2013), clarifying these relationships is critical for identifying at-risk populations and informing targeted prevention strategies.

While the adverse mental health effects of air pollution have been documented across the lifespan, adolescence may represent a window of heightened vulnerability, during which exposure can influence cognitive and emotional functioning (Baker et al., 2025; Casey et al., 2008; Herting et al., 2024). Several studies have found a link between air pollution exposure and the presence of depressive and anxiety symptoms in childhood and adolescence (Lin et al., 2026; Zundel et al., 2025). In addition, air pollution exposure has also been associated with psychotic-like experiences (PLEs), such as hearing voices, mild paranoia, or magical thinking (Bradley et al., 2024; Newbury et al., 2024). Of note, these early-emerging symptoms have been shown to predict a range of later adverse psychiatric and physical health problems (Lindgren et al., 2022; Lu et al., 2024; Petito et al., 2020; Sun et al., 2026), underscoring their importance as early indicators of vulnerability. Critically, examining trajectories of these symptoms, rather than cross-sectional levels, provides insight into their developmental course, including patterns of persistence, escalation or remission, which differentiate risk for later psychiatric and functional outcomes and provide predictive information beyond single time-point measures (e.g., (Tseliou et al., 2024)). However, longitudinal work studies examining the role of air pollution in shaping mental health trajectories during adolescence remain limited, particularly in population-based samples. Understanding how air pollution influences symptom trajectories during this period is essential for identifying early risk processes and informing prevention.

The link between air pollution exposure and mental health outcomes is unlikely to reflect environmental influences alone. Recent findings suggest that genetic liability may modulate individual sensitivity to air pollution to ultimately shape neurodevelopment (Li et al., 2023a, 2023b). Consistent with this notion, a large UK Biobank study showed that long-term exposure to air pollution was associated with higher schizophrenia risk, with the strongest effects observed in individuals with elevated genetic risk for schizophrenia (Liu et al., 2024). Similarly, exposure to higher levels of air pollution has been linked to increased risk of depression, particularly among individuals with higher PRS for major depressive disorder (Fu et al., 2022). Together, these studies suggest that the association between air pollution and mental health may be amplified by underlying genetic vulnerability. However, it remains unclear whether the interplay between pollution and genetic liability extends to earlier developmental periods and shapes the emergence and longitudinal course of mental health symptoms during adolescence.

The current study used data from the Adolescent Brain Cognitive Development (ABCD) Study to characterize how childhood exposure (age 9–10 years) to particulate matter (PM2.5) and genetic risk for schizophrenia and major depressive disorder independently and jointly shape changes in internalizing symptoms and PLEs during adolescence. We chose to focus on PM2.5 exposure because it contributes substantially to the health burden of air pollution, given its small particle size that enables deep penetration into the respiratory system (Bhattarai et al., 2024). In addition, modelling studies indicate that climate change is likely to drive increases in PM2.5 concentrations globally in the coming years (Huang et al., 2021; Shen et al., 2017). Therefore, the present study underscores the importance of investigating the health impact of PM2.5, particularly among vulnerable populations. Further, we focused on internalizing symptoms because adolescence represents a key developmental period for the emergence and escalation of anxiety and depressive symptoms, while externalizing and attention problems typically have an earlier onset in childhood (Doering et al., 2022; Liu, 2004). We also included PLEs because they are a transdiagnostic marker of vulnerability, predicting a broad range of psychopathology later in life (Giocondo et al., 2021; Lindgren et al., 2022). We hypothesized that greater exposure to PM2.5 in late childhood would be associated with greater increases in internalising symptoms and slower decrease in PLEs over time. We also hypothesized that higher genetic risk for schizophrenia and major depressive disorder would be independently associated with more adverse symptom trajectories as well as amplify the effects of PM2.5 exposure, such that associations between PM2.5 and psychopathology would be strongest among individuals with the highest genetic risk.

## Methods

### Participants

We used data from the ABCD Study (release 6.0; https://abcdstudy.org/), an ongoing longitudinal study on children recruited at the age of nine to ten from 21 sites across the United States to comprehensively characterize psychological and neurobiological development from early adolescence to young adulthood (Garavan et al., 2018). The present study used data from baseline through to the 6-year follow up. The final sample for the main analysis consisted of 10,620 participants who had complete data for air pollution and covariates at baseline as well as data on internalizing and/or PLEs at a minimum of one time point. The sample size and sociodemographic characteristics at each time point can be found in Table 1.

**Table 1.** Sociodemographic characteristics of the sample.

|  | 1-year | 2-year | 3-year | 4-year | 5-year | 6-year |
| --- | --- | --- | --- | --- | --- | --- |
| Sample size n | 10,620 | 10,372 | 9,879 | 9,156 | 8,398 | 1,215 |
| Sex |  |  |  |  |  |  |
| Male n | 5,572 | 5,457 | 5,202 | 4,822 | 4,407 | 644 |
| Female n | 5,078 | 4,915 | 4,677 | 4,334 | 3,991 | 571 |
| Age, years m (SD) | 10.97<br>(0.64) | 12.07<br>(0.67) | 12.96<br>(0.65) | 14.17<br>(0.71) | 15.08<br>(0.67) | 16.06<br>(0.64) |
| Internalizing symptoms | 1.74<br>(2.11) | 1.81<br>(2.22) | 2.03<br>(2.38) | 2.40<br>(2.66) | 2.78<br>(2.84) | 2.90<br>(2.86) |
| Psychotic like experiences | 4.58<br>(9.37) | 3.57<br>(7.80) | 2.87<br>(6.53) | 2.86<br>(6.96) | 2.50<br>(6.30) | 2.06<br>(5.79) |
| Combined Household Income |  |  |  |  |  |  |
| 1 % | 12.32 | 11.93 | 11.14 | 9.55 | 8.55 | 7.87 |
| 2 % | 12.86 | 12.77 | 11.68 | 11.26 | 10.71 | 12.75 |
| 3 % | 12.15 | 13.22 | 12.39 | 11.17 | 10.65 | 8.68 |
| 4 % | 13.20 | 13.85 | 14.09 | 12.76 | 12.04 | 13.74 |
| 5 % | 30.11 | 33.84 | 35.06 | 36.48 | 37.00 | 38.61 |
| 6 % | 12.06 | 14.39 | 15.62 | 18.79 | 21.05 | 18.35 |
| NA % | 7.30 | 0 | 0 | 0 | 0 | 0 |
| 2016 annual PM2.5: concentration (lg/m3) at baseline | 7.65<br>(1.56) | NA | NA | NA | NA | NA |

m: mean; NA: not applicable; PM2.5: particulate matter $\leq 2.5 \mu\text{m}$ ; SD: standard deviation. Internalizing symptoms are calculated as the Brief Problem Monitor (BPM) sum score. Psychotic-like experiences are calculated as the total severity score of the Prodromal Questionnaire–Brief Child Version (PQ-BC).

### Air pollution exposure measures

Details on the collection of residential addresses and linkage to one-year annual average ambient PM2.5 have been previously described (Fan et al., 2021). Briefly, the primary address at the baseline study visit was geocoded by the DAIRC using the Google map API to generate latitude and longitude (Google Maps Platform Documentation, 2019). Daily concentrations of PM2.5 were estimated at a 1-km² resolution across the United States using hybrid spatiotemporal models that integrate satellite-based aerosol optical depth, land-use regression and chemical transport models (Di et al., 2019). Their estimates were then averaged over the 2016 calendar year, when the children were aged 9-10 years and assigned to the geocoded primary residential address at the baseline ABCD study visit. The 2016 calendar year was chosen to correspond with the onset of enrolment in the ABCD study. See Supplementary Material for details.

### Computation of the polygenic risk score for schizophrenia and major depressive disorder

Detailed information about genotyping in ABCD has been previously described (Fan et al., 2023; Uban et al., 2018). Briefly, we extracted 7,989,528 genetic variants with minor allele frequency >0.01, SNP-level call rate >0.98 and Hardy–Weinberg equilibrium p >1e-10 based on the imputed genotyping data. We identified adolescents of European ancestry using the first 6 principal components with random forest models and unrelated individuals based on PC-AiR (Fan et al., 2023). After removing 36 adolescents with principal components outliers within the European ancestry population, we retained 4,470 unrelated adolescents of European ancestry. We calculated a PRS for schizophrenia (PRS-SCZ) based on a genome-wide association study (GWAS) of European ancestry, including 53,386 cases and 77,258 controls (Trubetskoy et al., 2022). PRS for major depressive disorder (PRS-MDD) was calculated based on GWAS of European ancestry, including 500,199 individuals (Howard et al., 2019).

Although these GWAS summary statistics are based on adults, previous research suggested a robust overlap between salient genetic factors in adolescents and adults (Alves et al., 2019). The single nucleotide polymorphism (SNP) weights for PRS construction were computed using PRS-CS software, a Bayesian scoring method which places a continuous shrinkage prior on SNP effect sizes (Ge et al., 2019). European samples from the 1000 Genomes Project Phase 3v5 reference panel were used to model linkage disequilibrium among variants. We then used PLINK to calculate polygenic scores for individuals in the target ABCD sample by summing all included variants weighted by the inferred posterior effect size for the effect allele.

### Mental health measures

We used youth-reported internalizing symptoms from the Brief Problem Monitor (BPM) (Pedersen et al., 2021), which was collected every 6 months starting at 6-months post baseline. Participants also completed the Prodromal Questionnaire–Brief Child Version (PQ-BC) annually starting at baseline, a 21-item measure validated in children using the ABCD Study sample (Karcher et al., 2020), assessing the presence of PLEs (i.e., unusual thought content and perceptual abnormalities) over the past month. All items were administered by trained research assistants. The total severity score was used as the measure of PLEs. Data from all available time points were included to model longitudinal symptom change.

### Statistical analysis

To examine associations between PM2.5 and mental health trajectories, we fitted linear mixed-effects models using the *lme4* package in R (version 4.4.2), with BPM internalizing subscale scores and PQ-BC total severity score as time-varying outcomes across ten and seven time points, respectively. Separate models were estimated for internalizing symptoms and PLEs. Given the sex differences in mental health trajectories in this sample (see Supplementary Material), sex-differences in both vulnerability to different types of psychopathology (Stainton et al., 2021; Stewart et al., 2022) and symptom trajectories during adolescence (Rakesh et al., 2025), analyses were stratified by sex.

We first examined how PM2.5 exposure, PRS-SCZ, PRS-MDD (included as predictors in separate models) were associated with change in symptoms over time by modelling an interaction term between the predictor and age (lower-order main effects were also included). Models adjusted for change associated with household and neighbourhood disadvantage—operationalized as household income and area deprivation index. We included random intercepts and random slopes for age at the participant level, as well as random intercepts for family and study site.

To examine the interplay between pollution exposure and genetic liability, we tested whether PRS-SCZ or PRS-MDD moderated the relationship between PM2.5 and mental health trajectories by modelling a 3-way interaction term between PM2.5, PRS, and age (lower-order interaction terms and main effects were also included). All models retained the same random-effects structure.

Analyses were corrected for multiple comparisons using a false discovery rate (FDR) of p < 0.05 (n=4 comparisons in each set of analysis). To ensure the robustness of our findings, we conducted sensitivity analyses to assess additional confounding effects of other pollutants, including nitrogen dioxide (NO₂) and ozone (O₃), as well as population density and distance to major roads.

## Results

### PM2.5 exposure is associated with mental health trajectories

In females, we found an association between PM2.5 exposure at ages 9-10 years and PLEs trajectories (Fig. 1a). Specifically, in females only, while PLEs decreased at all levels of PM2.5 exposure, higher PM2.5 exposure was associated with lower symptom decreases over time relative to lower PM2.5 exposure (β = 0.060, SE= 0.019, p-FDR = 0.005). There was no significant association between PM2.5 exposure and internalising symptoms trajectories in either sex (Tab. S.1).

**Fig. 1.**
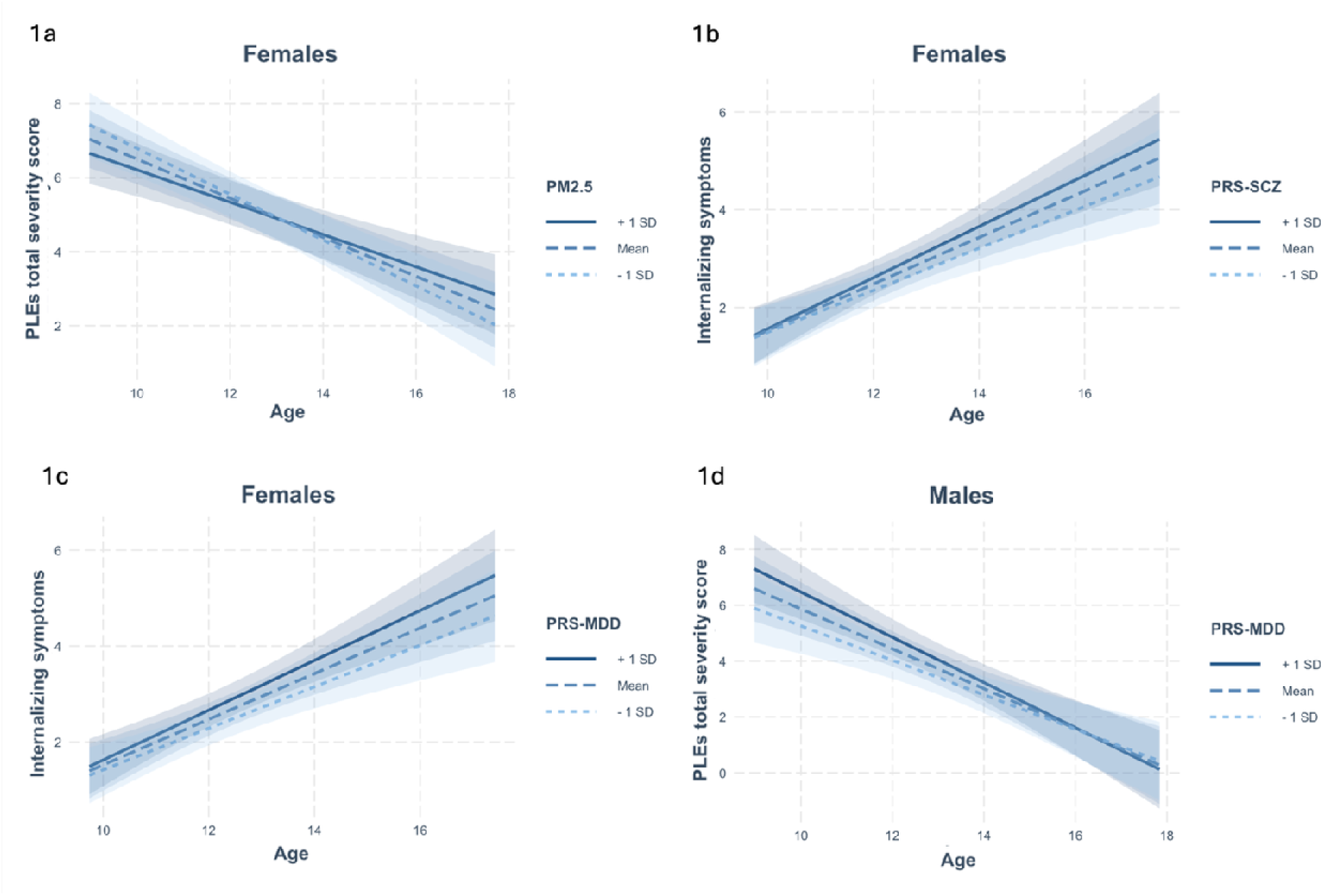
Sex-stratified associations between PM2.5, polygenic risk scores and mental health trajectories across adolescence. 1a. Association between PM2.5 exposure and psychotic-like experiences (PLEs) trajectories in females. 1b. Association between polygenic risk score for schizophrenia (PRS-SCZ) and internalising symptom trajectories in females. 1c. Association between polygenic risk score for major depressive disorder (PRS-MDD) and internalising symptom trajectories in females. 1d. Association between polygenic risk score for major depressive disorder (PRS-MDD) and PLEs trajectories in males. Lines represent predicted trajectories from linear mixed-effects models; shaded areas indicate 95% confidence intervals. Solid, dashed and dotted lines represent +1 SD, mean and −1 SD values of the moderator variables, respectively. Figures were generated using ggplot2 in R.

### Genetic risk is associated with mental health trajectories

We found that both PRS-SCZ and PRS-MDD were associated with internalising symptoms trajectories in females (Fig. 1b, 1c). Specifically, both higher PRS-SCZ and higher PRS-MDD were greater increases in internalising symptoms in females only (β = 0.124, SE= 0.041, p-FDR = 0.009; β = 0.368, SE= 0.132, p-FDR = 0.022).

Additionally, PRS-MDD was associated with PLEs trajectories in males (Fig. 1d), such that higher PRS-MDD was associated with a steeper rate of decrease of PLEs over time (β = −0.828, SE= 0.237, p-FDR = 0.001). No other associations were significant (Tab. S.1).

### Genetic risk moderates the association between PM2.5 exposure and mental health trajectories

PRS-MDD moderated the association between PM2.5 exposure and change in PLEs over time in females (β = 0.522, SE= 0.208, p-FDR = 0.048; Fig. 2) but not in males. Specifically, high PRS-MDD and high levels of PM2.5 exposure was associated with the greatest increases in PLEs over time, whereas lower exposure was associated with decreases in symptoms over time. No other 3-way interactions were significant (Tab. S.1).

**Fig 2.**
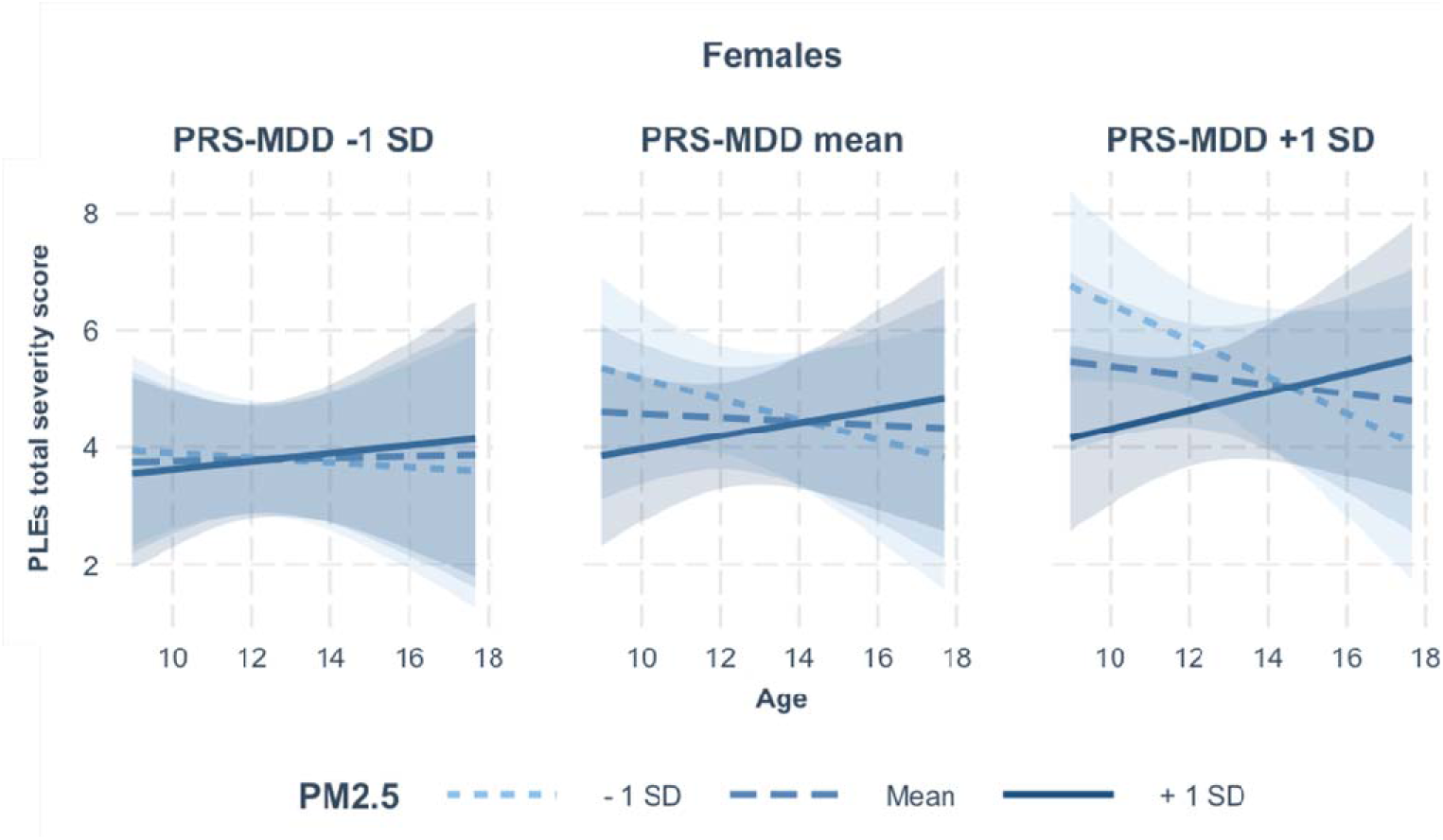
Associations between PM2.5, polygenic risk score for major depressive disorder (PRS-MDD) and psychotic-like experience (PLEs) trajectories in females. Lines represent predicted trajectories from linear mixed-effects models; shaded areas indicate 95% confidence intervals. Solid, dashed and dotted lines represent +1 SD, mean and −1 SD values of the moderator variables, respectively. Figures were generated using ggplot2 in R.

### Sensitivity analysis

Results remained significant in models that adjusted for additional covariates, including levels of other pollutants, population density and distance to roads (Tab. S.2).

## Discussion

In this longitudinal study using a large populational adolescent sample, we found evidence of sex-specific associations between childhood pollution exposure (PM2.5), genetic risk for mental health problems and trajectories of internalizing symptoms and PLEs. In females, higher pollution exposure at ages 9–10 years was associated with a slower decline in PLEs over time, suggesting greater persistence of PLEs during adolescence. In addition, both genetic risk for both SCZ and MDD were associated with increases in internalizing symptoms over time in females. We also provide novel evidence of a gene–environment interaction in females, such that the greatest increases in PLEs were observed in those with a combination of high genetic risk and high pollution exposure. This indicates that genetic susceptibility to major depressive disorder may confer risk for persistent PLEs in the context of higher PM2.5 exposure.

Our study shows that higher childhood PM2.5 exposure is associated with a slower decline in PLEs over time in females. This finding aligns with and extends a growing literature linking PM2.5 to adverse mental health outcomes (Khan et al., 2019; Liu et al., 2023), by showing that childhood pollution also influences the longitudinal course of PLEs beyond cross-sectional associations. One possible interpretation is that childhood exposure to PM2.5 may influence the neurodevelopmental processes implicated in the pathophysiology of PLEs, including salience processing, sensory integration and hypersensitivity to reward (Chun et al., 2020; Ermel et al., 2019; Sperandio et al., 2023). PM2.5 exposure has been linked to alterations in brain regions involved in these processes, including the insula, anterior cingulate cortex, prefrontal cortex and striatum (de Jesus et al., 2025), which are more directly implicated in the pathophysiology of PLEs. Disruption of the maturation of these systems during adolescence may contribute to the persistence of PLEs by influencing the developmental decline in unusual perceptual experiences and aberrant salience attribution typically observed during this period. Importantly, we found this association only in females, which suggests that sex-specific factors may modulate susceptibility to PM2.5 exposure. Differences in immune function have been proposed as potential mechanisms underlying sex differences in the relationship between environmental exposures and mental health outcomes (Zundel et al., 2025). Furthermore, evidence from animal studies indicates that exposure to traffic-related air pollution during early development can alter inflammatory responses and neurodevelopment in a sex-dependent manner (Patten et al., 2020). Taken together, these findings suggest that sex differences in inflammatory and neurodevelopmental processes may underlie the female-specific association between childhood PM2.5 exposure and the longitudinal course of PLEs observed in the present study. Our longitudinal findings stand in contrast to a previous investigation using the same cohort which reported an association between number of days with ambient PM2.5 levels above EPA standards and levels of internalizing symptoms one year later (Smolker et al., 2024). Differences in exposure metrics, outcome measures, and follow-up duration may account for these divergent findings. Whereas Smolker et al. focused on short-term associations with internalizing symptoms, our study examined longer-term trajectories of internalizing symptoms and PLEs from early to mid-adolescence.

PRS-SCZ and PRS-MDD were associated with increases in internalizing symptoms over time in females. This finding is consistent with the transdiagnostic nature of polygenic risk scores, with different genetic influences converging to shape internalizing symptom trajectories (Crouse et al., 2021; Kwong et al., 2021; Lussier et al., 2020). The presence of these associations only in females suggests that sex-related biological factors may modulate the phenotypic expression of genetic risk for schizophrenia and major depressive disorder, potentially through differences in hormonal regulation, emotion processing and stress responsivity across adolescence. Future studies are needed to clarify the mechanisms through which genetic risk contributes to sex-specific patterns of internalizing symptom development. Unexpectedly, in males, higher PRS-MDD was associated with a steeper decline in PLEs over time, while no association was observed with internalizing symptoms. This finding is challenging to interpret and appears somewhat inconsistent with previous evidence suggesting shared genetic influences between major depressive disorder and PLEs. It is possible that this association reflects sex-specific developmental processes, distinct manifestations of genetic liability across adolescence or other unmeasured factors. However, given the unexpected direction of the effect and the limited existing literature on the relationship between genetic liability for depression and longitudinal PLE trajectories, this finding should be interpreted cautiously. Replication in independent longitudinal cohorts will be important to establish its robustness and clarify its underlying mechanisms.

In line with our hypothesis, we observed multiplicative effects of genetic risk and air pollution exposure. PRS-MDD moderated the association between air pollution exposure and PLEs trajectories, with the greatest increase in PLEs observed in those with high PRS-MDD and high levels of PM2.5 exposure, whereas lower exposure was associated with decreases in symptoms over time. These findings are consistent with the diathesis–stress model, which describes how genetic liability may increase vulnerability to the adverse effects of environmental exposures (Monroe and Simons, 1991). Within this framework, PM2.5 may represent an environmental stressor that exacerbates the effects of genetic susceptibility to psychopathology. Importantly, this finding complements the observed association between PM2.5 exposure and slower decline in PLEs in females by suggesting that susceptibility to PM2.5 exposure may vary according to underlying genetic liability. Genetic risk for major depressive disorder has been associated with increased risk of PLEs in adolescence (Pain et al., 2018). In addition, a study conducted in a large cohort of the UK Biobank demonstrated that the combined impact of PM2.5 with high genetic susceptibility significantly amplified the risk of major depressive disorder in adults (Pan et al., 2025). Recent evidence highlights that environmental influences on youth mental health may operate through dynamic interactions with genetic and neurodevelopmental factors, with biological vulnerability acting as both a mediator and a moderator of associations between the environment and mental health in children and adolescents (Whittle et al., 2024). In line with this, higher PM2.5 exposure and higher polygenic risk for depression have been shown to jointly influence brain connectivity within networks involved in stress processing and emotional regulation (Li et al., 2021), suggesting that PM2.5 exposure may affect brain networks implicated in PLEs. Importantly, we found that PRS-MDD moderated the association between PM2.5 exposure and PLEs trajectories only in females. One possible explanation is that, in females, PM2.5 exposure may contribute to neuroinflammatory processes via peripheral immune activation and microglial sensitisation, leading to increased oxidative stress and altered neuronal integrity (Brockmeyer and D’Angiulli, 2016; Zhang et al., 2024). Within this context, genetic risk for major depressive disorder may further increase sensitivity to these processes, thus increasing vulnerability to disruptions in brain netowrks implicated in PLEs. Given evidence for sex differences in immune and inflammatory processes (Zundel et al., 2025), these mechanisms may be particularly relevant in females. Overall, these findings highlight the importance of integrating genetic and environmental risk factors to better understand the biological mechanisms underlying the development and persistence of PLEs.

The study has several strengths, including a large nation-wide cohort with air pollution estimates with high spatiotemporal precision, the longitudinal design allowing assessment of symptom trajectories from late childhood to adolescence and the ability to adjust for demographic and socio-economic confounders. However, some limitations should also be acknowledged. First, the observational nature of the study precludes causal inference and residual confounding is likely. Second, a single measure of PM2.5 concentration at baseline was used to quantify PM2.5 exposure. As measures of lifetime cumulative exposure are not available in the ABCD dataset, we were not able to assess the role of lifetime PM2.5 exposure on the longitudinal course of mental health symptoms during adolescence. Future studies that incorporate these measures will allow us to identify specific windows of susceptibility and the short- or long-term impact of air pollution on mental health. Lastly, the dataset does not provide comprehensive measures of near-roadway pollutants such as carbon dioxide, carbon monoxide and hydrocarbons (World Health Organization, 2013). Although sensitivity analyses suggested that our findings were robust to adjustment for other pollutants, population density and distance to main roads, these measures do not fully capture traffic-related pollution. More refined exposure metrics, including near-roadway pollutants, will be necessary to disentangle their effects on adolescent mental health.

In conclusion, our findings indicate that childhood exposure to PM2.5, together with its interaction with genetic risk for major depressive disorder, is associated with variation in mental health trajectories across adolescence. These results suggest that environmental exposures and genetic risk do not act independently, but jointly shape developmental pathways relevant to emerging psychopathology, with effects that differ by sex. Improving air quality is a complex but tractable issue (Mudway et al., 2019) and therefore efforts to improve air quality and reduce exposure during sensitive periods of development may represent a meaningful population-level strategy to support adolescent mental health.

## Data Availability

Data used in this study are from the Adolescent Brain Cognitive Development (ABCD) Study. The ABCD Study data are available to qualified researchers through the ABCD data access platform upon approval of a data use agreement. Further information on data access procedures is available through the ABCD Study website (https://abcdstudy.org). The authors did not collect the data and do not have the authority to grant access to the data.

## Funding and Acknowledgments

D.R. acknowledges support from a Young Investigator Grant from the Brain & Behaviour Research Foundation (32908) and a New Investigator Research Grant from the UKRI Medical Research Council (MR/Z506667/1).

Data used in the preparation of this article were obtained from the ABCD Study (https://abcdstudy.org) held in the NBCD. This is a multisite, longitudinal study designed to recruit more than 10,000 children aged 9–10 years old and follow them over 10 years into early adulthood. The ABCD study is supported by the National Institutes of Health and additional federal partners under award numbers <u>U01DA041048</u>, <u>U01DA050989</u>, <u>U01DA051016</u>, <u>U01DA041022</u>, <u>U01DA051018</u>, <u>U01DA051037</u>, <u>U01DA050987</u>, <u>U01DA041174</u>, <u>U01DA041106</u>, <u>U01DA041117</u>, <u>U01DA041028</u>, <u>U01DA041134</u>, <u>U01DA050988</u>, <u>U01DA051039</u>, <u>U01DA041156</u>, <u>U01DA041025</u>, <u>U01DA041120</u>, <u>U01DA051038</u>, <u>U01DA041148</u>, <u>U01DA041093</u>, <u>U01DA041089</u>, <u>U24DA041123</u> and <u>U24DA041147</u>. A full list of supporters is available at https://abcdstudy.org/federal-partners.html. A listing of participating sites and a complete listing of the study investigators can be found at https://abcdstudy.org/consortium_members/. ABCD consortium investigators designed and implemented the study and/or provided data but did not necessarily participate in the analysis or writing of this report. The views expressed in this manuscript are those of the authors and do not necessarily reflect the official views of the National Institutes of Health, the Department of Health and Human Services, the US federal government or ABCD consortium investigators.

