## Supplementary Material for "The independent and joint effects of outdoor air pollution exposure and genetic risk on mental health trajectories during adolescence"

**Supplementary Methods**

**Supplementary Results**

**Supplementary Figures**

**Supplementary Tables**

**Supplementary Methods**

**Air pollution exposure measures**

Details on the collection of residential addresses and linkage to one-year annual average ambient particulate matter (PM₂.₅), nitrogen dioxide (NO₂), and ozone (O₃) have been previously described (Fan et al., 2021). Briefly, the address at which participants spent most of their time was obtained from the participant’s caregiver during the baseline study visit and geocoded by the DAIRC using the Google map API to generate latitude and longitude (Google Maps Platform Documentation, 2019). Daily concentrations of PM₂.₅, NO₂, and 8-hour maximum ground-level O₃ were estimated at a 1-km² resolution across the United States using hybrid spatiotemporal models that integrate satellite-based aerosol optical depth, land-use regression, and chemical transport models. (Di et al., 2019). Their estimates were then averaged over the 2016 calendar year, when the children were aged 9-10 years and assigned to the geocoded primary residential address at the baseline ABCD study visit. The 2016 calendar year was chosen to correspond with the onset of enrolment in the ABCD study. PM₂.₅ was reported in µg/m3, and O₃ and NO₂ were reported in parts per billion (ppb). Nationwide, the annual PM₂.₅, NO₂, and O₃ concentration estimates are highly accurate, with a cross-validated R2 of 0.89, 0.84, and 0.90, respectively (Di et al., 2020, 2019; Fan et al., 2021). The mean (SD) annual pollutants concentrations from the 2016 calendar year were as follows: PM₂.₅: 7.65 (1.56) µg/m3, NO₂: 19.19 (6.17) ppb and O₃: 41.65 (4.86) ppb. PM₂.₅ and NO₂ levels were below current Environmental Protection Agency guidelines (https://www.epa.gov/criteria-air-pollutants).

**Supplementary Results**

**Sex differences in mental health trajectories**

Internalizing symptoms showed a significant positive association with age (β = 0.073, SE = 0.009, p < 2 × 10⁻¹⁶), and this association varied by sex (age × sex: β = 0.336, SE = 0.013, p < 2 × 10⁻¹⁶), such that the age-related increase was more pronounced in females compared to males. In contrast, PLEs decreased significantly with age in both sexes (β = −0.822, SE = 0.021, p < 2 × 10⁻¹⁶), with a significant age × sex interaction (β = 0.366, SE = 0.031, p < 2 × 10⁻¹⁶), indicating that the decline was steeper in males than in females (Fig. S.1).

**Association between PM₂.₅ exposure and PLEs in females adjusted for the effects of nitrogen dioxide (NO₂), and ozone (O₃), population density and distance to roads (sensitivity analysis).**

The association between PM₂.₅ exposure at ages 9-10 years and PLEs trajectories in females remained significant after adjusting for nitrogen dioxide (NO₂), ozone (O₃), population density and distance to roads (β = 0.060, SE= 0.015, p-FDR < 0.001) (Fig. S.2).

**Association between PM₂.₅ exposure, PRS-MDD and PLEs trajectories in females adjusted for the effects of nitrogen dioxide (NO₂), and ozone (O₃), population density and distance to roads (sensitivity analysis).**

In females, after adjusting for NO₂, O₃, population density and distance to roads, PLEs trajectories were significantly associated with PM₂.₅ exposure at ages 9-10 years and PRS-MDD, with a significant interaction effect between PM₂.₅ exposure and genetic risk (β = 0.542, SE = 0.208, p-FDR = 0.036) (Fig. S.3).

**Supplementary Figures**

**Fig. S.1 Sex differences in mental health trajectories**

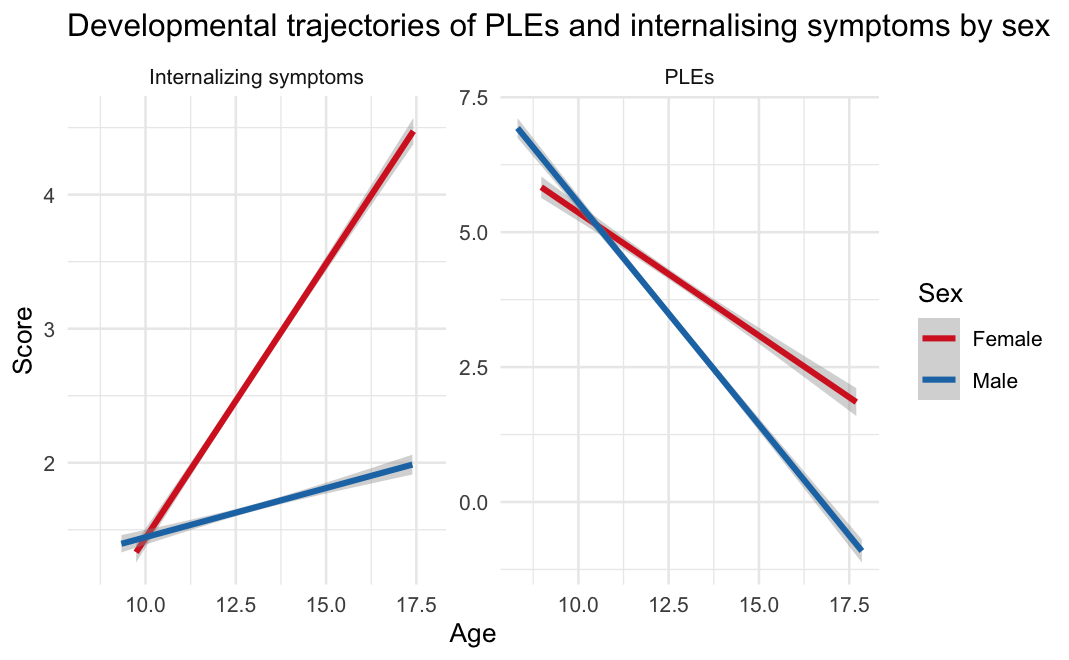
**Supplementary Tables**

**Tab. S.1 Associations of PM₂.₅ exposure and PRS-SCZ and PRS-MDD with internalizing symptoms and psychotic-like experiences in males and females**

|  | **β** | **SE** | **p-FDR** |
| --- | --- | --- | --- |
| **PM₂.₅** | | | |
| **Males** | | | |
| **Internalizing symptoms** | **-0.007** | 0.005 | 0.368 |
| **PLEs** | 0.026 | 0.015 | 0.261 |
| **Females** | | | |
| **Internalizing symptoms** | **-0.002** | **0.007** | 0.883 |
| **PLEs** | 0.060 | 0.019 | 0.005* |
| **PRS-SCZ** | | | |
| **Males** | | | |
| **Internalizing symptoms** | -0.009 | 0.031 | 0.900 |
| **PLEs** | -0.044 | 0.072 | 0.812 |
| **Females** | | | |
| **Internalizing symptoms** | 0.124 | 0.041 | 0.009* |
| **PLEs** | 0.001 | 0.094 | 0.992 |
| **PRS-MDD** | | | |
| **Males** | | | |
| **Internalizing symptoms** | **0.079** | **0.1006** | 0.713 |
| **PLEs** | 0.828 | 0.237 | 0.001* |
| **Females** | | | |
| **Internalizing symptoms** | 0.368 | 0.132 | 0.022* |
| **PLEs** | **-0.386** | 0.306 | 0.368 |
| **PM₂.₅ x PRS-SCZ** | | | |
| **Males** | | | |
| **Internalizing symptoms** | 0.001 | 0.019 | 0.992 |
| **PLEs** | 0.069 | 0.046 | 0.288 |
| **Females** | | | |
| **Internalizing symptoms** | -0.019 | 0.026 | 0.760 |
| **PLEs** | 0.056 | 0.062 | 0.620 |
| **PM₂.₅ x PRS-MDD** | | | |
| **Males** | | | |
| **Internalizing symptoms** | -0.067 | **0.067** | 0.570 |
| **PLEs** | -0.043 | **0.1592** | 0.900 |
| **Females** | | | |
| **Internalizing symptoms** | **0.058** | **0.090** | 0.775 |
| **PLEs** | 0.522 | 0.208 | 0.048* |

**Tab. S.2 Associations of PM₂.₅ exposure and PRS-MDD with psychotic-like experiences in males and females adjusted for additional covariates, including levels of other pollutants, population density and distance to roads (sensitivity analysis)**

|  | **β** | **SE** | **p-FDR** |
| --- | --- | --- | --- |
| **PM₂.₅** | | | |
| **Females** | | | |
| **PLEs** | 0.081 | 0.019 | 0.004* |
| **PM₂.₅ x PRS-MDD** | | | |
| **Females** | | | |
| **PLEs** | **0.539** | 0.207 | 0.037* |
